# Acceptability, Appropriateness and Feasibility of a Calibrated Obstetric Blood Collection Drape for Postpartum Haemorrhage Detection: A Qualitative Study in Two Tertiary-Care hospitals in Delhi, India

**DOI:** 10.64898/2026.08.11.26360170

**Authors:** Subhanwita Manna, Pratibha Dhiman, Tanica Lyngdoh, Hrishikesh Munshi, Suyesh Goel, Jyotsna Suri, Monika Gupta, Rajesh Kumari, Bindu Bajaj, Rajneesh Joshi, Pranjali V Dhume, Reema Mukherjee

## Abstract

**Background:** Postpartum haemorrhage (PPH) is the leading cause of maternal mortality globally. Accurate and objective blood loss measurement is essential for timely detection and management of PPH. However, visual estimation remains the predominant method of blood loss estimation in low- and middle-income country settings, despite it underestimating blood loss by 33-50%. In this context, a calibrated obstetric blood collection drape offers a practical, low-cost alternative. However, Evidence on acceptability, appropriateness and feasibility remains limited.

**Objective:** To explore the facilitators and barriers to the acceptability, appropriateness and feasibility of the routine implementation of a calibrated obstetric blood collection drape for postpartum haemorrhage (PPH) detection in two tertiary-care hospitals in Delhi, India.

**Methods:** This qualitative study used semi-structured interviews to explore healthcare providers’ experiences and perceptions regarding the acceptability, appropriateness and feasibility of using the drape in routine labour room practice. Qualitative data were analysed using thematic analysis.

**Results:** Eighteen healthcare providers participated in qualitative interviews. The calibrated drape was perceived as more reliable than visual estimation and easy to use, supporting confidence, quicker responses and clinical decision-making. It was also considered useful for early PPH detection and timely management. Key challenges included staff shortages and concerns about drape slipperiness and fastening mechanisms.

**Conclusions:** Findings suggest that the calibrated obstetric blood collection drape is perceived as acceptable, appropriate and feasible for routine use in high-volume tertiary-care hospitals in India. Wider implementation will require reliable supply chains, design refinements, and sustained training programmes, including for support staff.

## INTRODUCTION

Postpartum haemorrhage (PPH) remains the leading cause of maternal mortality worldwide, accounting for approximately 27% of all maternal deaths and an estimated 70,000 deaths annually, with the greatest burden occurring in LMICs (1,2). Despite substantial reductions in maternal mortality over recent decades (3,4), PPH continues to contribute significantly to maternal deaths and severe maternal morbidity in India (4,5).

Early recognition of excessive postpartum blood loss is critical for timely intervention and improved maternal outcomes (5,6). However, blood loss assessment in routine clinical practice is commonly based on visual assessment (5), which underestimates actual blood loss by 33-50% (7). Recognising this limitation, the World Health Organization (WHO) recommends routine objective measurement of postpartum blood loss to improve the detection and management of PPH (8).

Calibrated obstetric blood collection drapes provide a simple, low-cost, and objective method for measuring postpartum blood loss. These devices allow direct quantification of blood loss and can support earlier identification of women requiring escalation and management (7–9). Despite their demonstrated value, calibrated drapes have not been widely integrated into routine care in public healthcare settings in India.

Successful implementation of new technology depends not only on its effectiveness but also on its acceptability, feasibility, and appropriateness within routine clinical workflows (10–12).

Therefore, this study aimed to explore the facilitators and barriers to the acceptability, feasibility, and appropriateness of the routine implementation of a calibrated obstetric blood collection drape for postpartum haemorrhage (PPH) detection in two tertiary-care hospitals in Delhi, India

## Methods

### Study Design

The study used a qualitative approach, in particular, in-depth interviews to explore healthcare providers’ experiences and perceptions of using the calibrated drape in routine labour room practice in an open-ended way so that participants were given the opportunity to express a wide range of feelings and opinions on a topic asked. Prior to introducing the drape in the two study hospitals, healthcare providers received training on its use. Additional implementation support activities, including pre- and post-training assessments and on-the-job observations, were conducted to facilitate implementation and monitor adherence to the drape-use protocol. These activities were undertaken independently of the qualitative study and are reported in the Supplementary Material.

### Study Setting

The study was conducted in two government tertiary hospitals in Delhi, India. Both hospitals are high-volume referral centres providing comprehensive obstetric care and managing a large proportion of complicated pregnancies referred from peripheral facilities. The delivery volume and number of PPH cases are presented in Table 1. The study was carried out in two labour rooms of Hospital 1 and the single labour room of Hospital 2

**Table 1.** Delivery load and PPH burden at study hospitals, 2023-2024.

| Study site | Year | Total deliveries | Caesarean sections | Vaginal deliveries | Instrumental deliveries | PPH cases |
| --- | --- | --- | --- | --- | --- | --- |
| Hospital-1 | 2023 | 22,327 | 6,439 | 15,888 | 501 | 153 |
|  | 2024 | 17,617 | 5,595 | 12,022 | 283 | 118 |
| Hospital-2 | 2023 | 2,573 | 961 | 1,496 | 116 | 35 |
|  | 2024 | 2,656 | 1,034 | 1,539 | 83 | 32 |

### Training and implementation support

Training on the use of the calibrated drape was provided to all healthcare providers posted in the labour rooms of both hospitals. At the time of the study, 31 healthcare providers (18 residents and 13 nursing officers) were posted in the labour room of Hospital 1, while 27 healthcare providers (12 residents and 15 nursing officers) were posted in the labour room of Hospital 2.

Training was conducted by the research team, with technical support from the manufacturer’s representatives, to ensure Standardised and appropriate use of the calibrated obstetric blood collection drape. The training covered correct drape placement, timing of application, measurement and interpretation of blood loss, handling and disposal procedures, and infection prevention practices. The training consisted of one day interactive session that included presentations, live demonstrations of drape application and blood loss measurement, and supervised hands-on practice to familiarise participants with its use in routine clinical settings.

Following training, the calibrated obstetric drape was introduced into routine practice and used for all vaginal deliveries conducted in the participating labour rooms across both hospitals during a two-month implementation period. Healthcare providers therefore had direct experience using the drape before being considered for qualitative interviews. Detailed specifications of the drape are provided in Supplementary Table 1 and Supplementary Figure 1.

### Participant Selection and Recruitment

Participants were selected using purposive sampling because they were considered best informed to provide insights into the research questions. Participants comprised healthcare providers (including obstetric residents and nursing officers) who had routinely used the calibrated obstetric blood collection drape as part of clinical care, thereby providing experience-based insights into its acceptability, feasibility, and appropriateness. A total of 18 healthcare providers from both hospitals were interviewed, comprising 12 obstetric residents and 6 nursing officers. The final sample size was determined by thematic saturation, with recruitment continuing until no new themes or insights emerged from the interviews.

### Data Collection

A semi-structured interview guide was developed in accordance with the study objectives to explore participants’ perspectives on the facilitators and barriers to implementing the calibrated obstetric blood collection drape in routine clinical practice. The interview guide contained open-ended questions on experiences and challenges of the healthcare providers on implementing the use of drape, appropriateness and acceptability aspects (advantages over visual estimation, willingness to recommend to others and continue use, whether it improves blood loss assessment, effect on clinical decision-making, ease of use) and feasibility (ease of learning how to use and using correctly, workflow integration and perceived ease of routine implementation). Qualitative data were collected between September and November 2025. All interviews were conducted in English in private spaces within the hospital to ensure confidentiality. Interviews lasted approximately 20-35 minutes and were audio-recorded with participants’ written informed consent. The interviews were conducted by two members of the research team (SM and PD), who had prior training and experience in qualitative research. Participants were informed about the purpose of the study and the interviewers’ role before the interviews. The interviewers had no supervisory or managerial relationship with the participants.

### Data Quality control

All qualitative interviews were conducted using a standardised interview guide. Interviewers received training in qualitative interviewing, and regular discussions were held among the research team to ensure consistency throughout data collection.

### Data Management

Interviews were audio-recorded and transcribed verbatim. Transcripts were reviewed against original recordings for accuracy; all personal identifiers were removed, and participants were assigned unique identification codes corresponding to their professional role and study site. Transcripts, field notes, and reflexive memos were stored and managed using NVivo version 12 (QSR International, Melbourne, Australia).

### Data analysis

Qualitative data were analysed using an inductive thematic analysis approach following the framework described by Braun and Clarke (13). Transcripts were read repeatedly to familiarise the research team with the data. Two researchers independently coded a subset of transcripts; the coding framework was then refined through discussion and consensus. Codes were grouped into categories and developed into themes, which were reviewed for coherence and relevance to the study objectives. Representative quotations were selected to support the interpretation of findings.

## RESULTS

### Participants

We interviewed 18 healthcare providers comprising 12 obstetric residents (first-year: n=2; second-year: n=4; senior resident: n=6) and 6 nursing staff. Nursing staff had between 3 to 15 years of clinical experience in labour room settings, while residents had been working in obstetrics for 1-3 years depending on their training level. All interview participants had completed the standardised training and had used the calibrated drape routinely for approximately two months prior to interview. Among interview participants, 15 (83%) were female.

### Theme 1: Moving beyond guesswork to estimate blood loss

#### PPH requires early and accurate recognition

Participants consistently identified PPH as an important and commonly encountered challenge, and the need to be constantly vigilant to detect it in time to prevent mortality or further morbidity.

> *“It’s a nightmare for an obstetrician or a team leader or as a senior working in the labour room, everybody, every woman is delivering. The only thing that we have to be careful about is postpartum haemorrhage. I would say, like, an obstetric emergency. So, we need to be on our toes to manage that” [Resident 1, Hospital 1]*
>
> *“PPH is extremely serious and needs immediate attention. Identifying it late can lead to loss of life. It must be identified early, even by junior staff” [Resident 3, Hospital 1]*
>
> *“PPH is common here. Around 10 out of 100 patients have PPH because we handle many high-risk pregnancies. It is dangerous-if we underestimate blood loss, we can even lose the patient. So timely detection is very important” [Resident 6, Hospital 1]*
>
> *“Here we have lost one lady, maybe, when I was here for about four months, I guess it was because of PPH. Otherwise, PPH is taken very seriously in the labour room” [Nursing Officer 9, Hospital 2]*

#### Visual estimations are subjective

Despite recognizing the importance of timely PPH detection, participants consistently highlighted the limitations of visual blood loss estimation, describing it as subjective, inconsistent, and highly dependent on individual experience.

> *“The problem with visual estimation is that it depends on experience, and even experienced staff can be wrong. Blood looks like more or less depending on how it’s spread out. We needed something more objective” [Nursing Officer 6, Hospital 2].*
>
> *“It is a guesswork. Some people might underestimate the amount of bleeding, while others may overestimate it. There’s no Standardised or universal way to judge it, it really depends on the observer” [Resident 1, Hospital 1]*
>
> *“Blood mixed with amniotic fluid, spread on linen, it’s hard to judge. Sometimes if we underestimate, and by the time we realise it’s PPH, the patient is already in trouble, so we need to be very careful” [Resident 5, Hospital 1]*

### Theme 2: Experiences of using the Calibrated Drape

#### Easy to learn and integrate into routine practice

Training was considered simple and easy to understand, with several participants noting that even junior staff and postgraduate trainees could learn its use quickly.

> *“Training was very helpful and easy to understand. It is a simple method and even junior staff can easily learn and use it” [Nursing Officer 5, Hospital 1]*
>
> *“I initially found tying it cumbersome, but after using it for a few deliveries it became comfortable” [Resident 1, Hospital 1]*
>
> *“Our work became much easier after we started using these drapes” [Nursing Officer 3, Hospital 2]”*
>
> *“It fitted quite easily into our daily work and now feels like a normal part of delivery care” [Resident 3, Hospital 2]*
>
> *“It blended smoothly into the workflow after the initial few days” [Resident 6, Hospital 1]*
>
> *“Regular use and familiarity made it easy, once it became habitual it did not feel like an additional task “[Resident 5, Hospital 2]*

#### Confident about estimating blood loss

The participants felt confident that using the drape helped them accurately estimate the blood loss

> *“With the drape I could actually quantify how much blood was lost” [Resident 1, Hospital 2]*
>
> *“We can quantify it way better than before” [Nursing officer 1, Hospital 1]*
>
> *“We felt confident and accurate in estimating blood loss” [Resident 4, Hospital 1]*
>
> **“The calibrated markings made it easy to measure blood loss accurately and encouraged regular use” [Resident 3, Hospital 1]**

#### Perceived acceptability among women

Participants further reported that the drape was generally well tolerated by women. Although some providers noted occasional mild discomfort related to the material or tying mechanism, these concerns were not considered sufficient to limit routine use.

> *“No major adverse reactions were observed, but some patients felt discomfort because the drape sticks to their skin and does not absorb sweat” [Nursing Officer 7, Hospital 1]*

### Theme 3: Objective blood loss measurement enabled earlier and more confident clinical decision-making

#### Earlier recognition and timely intervention

Most of the participants reported that objective quantification of blood loss enabled faster clinical response. Accurate measurement supported earlier recognition of excessive bleeding, prompt escalation to senior staff, and timely initiation of treatment. Several participants noted that quantified estimates facilitated clearer communication with the blood bank regarding transfusion requirements.

> *“Sometimes we intervene a little earlier than before. Even if it is four hundred millilitres, we become alert and prepare for possible postpartum haemorrhage” [Resident 6, Hospital 1]*
>
> *“It leads to earlier intervention; otherwise, we would wait until two or three pads were soaked” [Nursing Officer 3, Hospital 2]*
>
> *“It gives an exact measurement of how much blood the patient is losing. Based on that, we can start the intervention as early as possible” [Resident5, Hospital 1]*

#### Objective measurement supported improved communication and clinical decisions

Objective measurement changed the way clinicians recognized, communicated about, and managed postpartum haemorrhage. It facilitated quicker clinical decisions.

> *“With the drape I could tell the blood bank exactly how much blood the patient had lost and what number of packed cells were needed” [Resident 5, Hospital 1]*
>
> *“Once blood loss is quantified, we can move quickly to management. It increased both speed and confidence in decision making” [Resident 2, Hospital 2]*
>
> *“Everyone was on the same page, and we could immediately see the exact amount of blood loss. It helped us take quick decisions and make timely interventions” [Resident 2, Hospital 2]*
>
> *“In one case with thrombocytopenia we anticipated heavy bleeding but the drape showed only one hundred millilitres and prevented unnecessary interventions” [Resident 4, Hospital 2]*

### Theme 4: Making routine implementation of calibrated drape work

#### Operational Challenges

Participants reported that, consistent drape use was challenging during periods of high patient volume or staff shortages.

> *“During heavy patient load, especially in busy shifts, it becomes difficult to use it for every patient. We are not used to using it, so we forget” [Nursing Officer 7, Hospital 1]*
>
> *“So, I have been familiar with this method since I joined and these drapes were effective. But sometimes, due to staff shortage or difficult situations, they were not used correctly, which reduces their effectiveness” [Nursing Officer 8, Hospital 1]*

#### Design refinements can improve usability

Participants identified certain design-related, challenges that could affect its sustained implementation. The most commonly reported concerns related to the drape material, which many participants described as slippery, short ties, need for bolder calibration, improvement in quality of nozzle to prevent accidental spillage.

> ***“**The drape is generally fine, but it is a bit slippery. Sometimes when the patient is very exhausted and sweats a lot, they start slipping, which makes it difficult for us” [Resident 1, Hospital 2]*
>
> *“The cloth is a bit slippery, and it makes it difficult to handle the patient. Sometimes the patient sweats a lot, and it slips” [Resident 4, Hospital 2]*
>
> *“The ties are short, making it difficult to secure on larger patients. It should be available in multiple sizes with longer straps” [Nursing Officer 9, Hospital 2]*
>
> *“The nozzle should have a lock so it does not open by mistake” [Nursing Officer 9, Hospital 1]*
>
> *“The calibration markings could be bolder for quick reading” [Nursing Officer 7, Hospital 1]*

#### Support for wider implementation

Despite these limitations, participants strongly supported wider implementation of the drape and consistently recommended its use in other healthcare facilities. Participants suggested that successful scale-up would require reliable procurement and uninterrupted supply, periodic refresher training for labour room staff, and minor design modifications, particularly improvements to the fabric, fastening mechanism, and drainage nozzle.

> *“Definitely, I will recommend it to others because it gives exact measurement of blood loss and helps diagnose PPH early and manage it timely” [Resident 3, Hospital 2]*
>
> *“I would definitely recommend it. In my five to six years of service, I haven’t seen anything that provides this level of accuracy” [Nursing Officer 9, Hospital 2]*
>
> *“Training should be given to staff who actually apply the drape, like orderlies and ward workers” [Resident 9, Hospital 1]*

## Discussion

This study explored healthcare providers’ experiences with implementation of a calibrated obstetric blood collection drape in two high volume tertiary care government hospitals in Delhi. The findings suggest that healthcare providers perceived the calibrated obstetric drape to be an acceptable, appropriate and feasible tool for routine assessment of PPH. Participants consistently described visual estimation of blood loss as subjective and unreliable. In contrast, the calibrated drape was perceived to provide a more objective assessment of blood loss supported earlier recognition of excessive bleeding and increased confidence in clinical decision-making.

Participants perceived the calibrated drape to be appropriate for routine labour room practice because it provided an objective method of blood loss assessment while visual estimation was considered subjective and inconsistent. Participants described visual estimation as “guesswork” that depended heavily on individual experience and could be influenced by factors such as blood mixing with amniotic fluid or absorption into linen. Against this background, the calibrated drape was viewed as offering a more Standardised approach to measuring blood loss. These perceptions are consistent with WHO and FIGO recommendations advocating objective measurement of postpartum blood loss (8,9) and support current efforts to incorporate quantified blood loss assessment into routine PPH detection.

Healthcare providers also perceived the drape to be acceptable in routine practice. Healthcare providers described it as simple to learn and reported that it became easier to use with repeated practice. Although some providers initially experienced difficulties with drape placement, these challenges diminished over time as the drape became integrated into routine workflow. Participants also reported that women generally accepted the drape, with only occasional concerns regarding discomfort related to the material.

The findings suggest that routine implementation of the calibrated obstetric drape is feasible within high-volume tertiary-care labour rooms, although successful implementation depends on addressing several operational considerations. Participants generally described the drape as easy to learn and reported that it became easier to use with repeated practice, indicating that it could be incorporated into routine labour room workflows. However, they also identified several practical challenges that could affect sustained implementation, including staff shortages during busy shifts, high patient volume, and occasional difficulties in remembering to use the drape consistently. Participants also highlighted design-related concerns, such as the slippery nature of the material, short fastening ties, and the need for improvements to the drainage nozzle and calibration markings. Importantly, these issues were viewed as modifiable implementation barriers rather than reasons to discontinue use of the drape. Participants consistently recommended reliable procurement, uninterrupted availability of drapes, periodic refresher training, and minor design refinements to support long-term routine implementation.

Our findings are consistent with previous implementation studies which have reported that simplicity, ease of use, and perceived clinical usefulness are important determinants of successful adoption of blood-loss measurement devices in routine obstetric practice (10–12). Similar implementation challenges emerged in the E-MOTIVE process evaluations in Kenya and Nigeria, where workload, staff constraints, and resource limitations were consistently cited as key barriers to reliable implementation (10,11). Additionally, FIGO 2025 explicitly acknowledges that device availability, supply costs, and environmental sustainability remain recurring implementation hurdles across LMIC settings (9).

The burden of PPH in Indian tertiary centres is substantial, underscoring the urgent need of scalable detection interventions (4,14–16). However, successful implementation will require attention to procurement, supply-chain management, and long-term sustainability (9,11). The need for affordable, sustainable, and context-appropriate blood-loss measurement devices is therefore both timely and critical for Indian public health settings (8,17).

### Strength and Limitations

A major strength of this study is that it explored healthcare providers’ experiences after the calibrated obstetric drape had been implemented under routine clinical conditions in two high-volume government tertiary hospitals. Participants had direct experience using the drape in everyday labour room practice, allowing the findings to reflect real-world implementation rather than hypothetical perceptions. The inclusion of both obstetric residents and nursing officers provided perspectives from different professional groups involved in postpartum care. This study also has several limitations. First, it was conducted in only two tertiary-care hospitals in Delhi, and the findings may not be applicable to lower-level facilities or settings with different staffing patterns and patient volumes. Second, healthcare providers had used the drape for approximately two months before the interviews; therefore, the findings primarily reflect early implementation experiences rather than long-term adoption and sustainability. Third, only healthcare providers were interviewed. Women’s perspectives on the acceptability and comfort of the drape were not explored.

## Conclusion

WHO guidelines have firmly shifted the global standard toward routine objective blood loss measurement for all births. Our study provides the operational evidence needed to support this shift in the Indian context. To move from “acceptability” to “standard of care,” health systems must ensure a robust supply chain, integrate drape-based measurement into routine labour room checklists, and extend training to support staff (orderlies), who are often the ones physically preparing the delivery environment. The study highlighted the perception of the healthcare providers towards the obstetric drape as an acceptable, appropriate, and feasible tool for routine postpartum blood loss assessment, however, the operational and system level challenges also identified, and were viewed as addressable through ongoing training, reliable supply systems, and minor product modifications. Future research should evaluate implementation in lower-level health facilities and examine the impact of routine drape use on clinical outcomes and referral practices.

## Declarations

## Ethical considerations

Ethical approval was obtained from the Institutional Vardhman Mahavir Medical College & Safdurjung Hospital, New Delhi (Ref. No-IEC/VMMC/SJH/CERT/2025-may/39; dated-26/05/2025]). Written informed consent was obtained from all participants. Each participant was informed of their right to decline or withdraw from the study at any point before or during data collection, with or without providing a reason. To ensure complete anonymization, numerical identifiers were used instead of names for all data. The research findings were disseminated through peer-reviewed publications and conference proceedings.

## Consent for publication

Not Applicable

## Data availability statement

The datasets generated and analysed during the current study are not publicly available due to ethical restrictions on sharing participant-level qualitative data but are available from the corresponding author on reasonable request and with appropriate ethics approvals.

## Author contributions

Subhanwita Manna and Pratibha Dhiman contributed equally as joint first authors and led data collection, qualitative analysis, and manuscript preparation. Reema Mukherjee and Tanica Lyngdoh provided overall oversight and methodological guidance. Hrishikesh Munshi reviewed and revised the protocol of the study on which this manuscript is based. Suyesh Goel, Jyotsna Suri, Hrishikesh Munshi, Pranjali V Dhume, Monika Gupta, Rajesh Kumari, Bindu Bajaj, and Rajneesh Joshi contributed to critical review of the manuscript and provided subject-matter inputs. All authors read and approved the final manuscript.

## Acknowledgement

The authors gratefully acknowledge the support of Medicare Hygiene Limited for providing the necessary materials for this study. We also extend our thanks to Women’s Collective Forum for logistical coordination.

We would like to express our sincere appreciation to Dr RM Pandey, Dr Nomita Chandiok, Dr Sheela Mane,Dr Sudipto Roy,Dr. Latha Venkatesan for their expert guidance and valuable suggestions throughout the study.

## Supporting Information

Supplementary Table 5. Standards for Reporting Qualitative Research (SRQR) checklist

## REFERENCES

1. Say L, Chou D, Gemmill A, et al. Global causes of maternal death: a WHO systematic analysis. Lancet Glob Health. 2014;2(6): e323–e333. doi:10.1016/S2214-109X(14)70227-X

2. World Health Organization. Roadmap to Combat Postpartum Haemorrhage 2023-2030. Geneva: WHO; 2023.

3. World Health Organization, United Nations Children’s Fund (UNICEF), United Nations Population Fund (UNFPA), World Bank Group, United Nations Department of Economic and Social Affairs, Population Division. Trends in maternal mortality 2000 to 2023: estimates by WHO, UNICEF, UNFPA, World Bank Group and UNDESA/Population Division. Geneva: World Health Organization; 2025.

4. Meh C, Sharma A, Ram U, et al. Trends in maternal mortality in India over two decades in nationally representative surveys. BJOG. 2022;129(4):550–561.

5. Bienstock JL, Eke AC, Hueppchen NA. Postpartum hemorrhage. N Engl J Med. 2021; 384:1635–1645. doi:10.1056/NEJMra1513247

6. Wormer KC, Jamil RT, Bryant SB. Acute Postpartum Hemorrhage. In: StatPearls. Treasure Island (FL): StatPearls Publishing; updated 2024.

7. Patel A, Goudar SS, Geller SE, Kodkany BS, Edlavitch SA, Wagh K, et al. Drape estimation vs. visual assessment for estimating postpartum hemorrhage. Int J Gynaecol Obstet. 2006;93(3):220–224. doi: 10.1016/j.ijgo.2006.02.014

8. World Health Organization. WHO Recommendations on the Assessment of Postpartum Blood Loss and Use of a Treatment Bundle for Postpartum Haemorrhage. Geneva: WHO; 2023. ISBN 978-92-4-507497-1.

9. Begum J, Duvekot JJ, Middleton P, et al.; FIGO. FIGO recommendations on objective measurement of blood loss after birth for early detection of postpartum hemorrhage. Int J Gynaecol Obstet. 2025. doi:10.1002/ijgo.70523

10. Bohren MA, Akter S, Moran NF, Gallos ID, Devall AJ, Middleton L, et al. Early detection and a treatment bundle strategy for postpartum haemorrhage: a mixed-methods process evaluation. Lancet Glob Health. 2025. doi:10.1016/S2214-109X(24)00454-6

11. Forbes G, Akter S, Miller S, Galadanci H, Qureshi Z, Fawcus S, et al. Factors influencing postpartum haemorrhage detection and management and the implementation of a new postpartum haemorrhage care bundle (E-MOTIVE) in Kenya, Nigeria, and South Africa. Implement Sci. 2023;18(1):1. doi:10.1186/s13012-022-01253-0.

12. Esau J, Morris T, Muller C, Els C, de Waard L. Two postpartum blood collection devices: the Brass-V Drape and Materna Well Tray-as experienced by birth attendants and birthing women-a questionnaire-based randomised study. Obstet Gynecol Int.2024; 2024:6605833. doi:10.1155/2024/6605833

13. Braun V, Clarke V. Using thematic analysis in psychology. Qual Res Psychol. 2006;3(2):77–101. doi:10.1191/1478088706qp063oa

14. Singh D, Rana D, Verma A. A prospective observational study on incidence, etiology, maternal risk factors, management and outcomes of postpartum haemorrhage in a tertiary care hospital. Int J Health Sci Res. 2026;16(1):101–109. doi:10.52403/ijhsr.20260113

15. Office of the Registrar General, India. Special Bulletin on Maternal Mortality in India 2021-23. New Delhi: Ministry of Home Affairs, Government of India; 2025.

16. World Health Organization. Consolidated Guidelines for the Prevention, Diagnosis and Treatment of Postpartum Haemorrhage. Geneva: WHO; 2025. ISBN 978-92-4-011611-5.

17. Binyamin Y, Orbach-Zinger S, Heesen M. Current strategies for the diagnosis and management of postpartum hemorrhage: a focused review of four Cochrane systematic reviews from 2024 and 2025. Int J Obstet Anesth. 2025; 63:104692. doi: 10.1016/j.ijoa.2025.104692

