## supplementary material for "Acceptability, Appropriateness and Feasibility of a Calibrated Obstetric Blood Collection Drape for Postpartum Haemorrhage Detection: A Qualitative Study in Two Tertiary-Care hospitals in Delhi, India"

**Supplementary Table 1.** Specifications of the calibrated obstetric blood collection drape used in the study

| Component | Description | Length | Width |
|---|---|---|---|
| Back film | Blue; low-density polyethylene (LDPE), 210 gauge | 60–65 cm | 90–95 cm |
| Funnel | Transparent; low-density polyethylene (LDPE), 250 gauge | 70–75 cm | 55–60 cm |
| Ties/strips | Blue; SMS non-woven fabric) | 20–25 cm | 5 cm |
| Outlet | Transparent; polypropylene | — | — |
| Strip wire | White; composite metal and plastic (funnel support) | — | — |
| Sterilisation | Ethylene oxide (EO) sterile | — | — |
| Sealing | Heat sealing | — | — |
| Packing material | White; medical-grade paper | 35 cm | 25 cm |
| Measurement capacity | 50-1,000 mL in 50 mL increments;<br>1,000-2,000 mL in 100 mL increments;<br>2,000-3,000 mL in 200 mL increments | — | — |

*EO, ethylene oxide; LDPE, low-density polyethylene; SMS, spunlace–meltblown–spunlace. Dimensions represent manufacturer-stated ranges. Dashes (—) indicate that the dimension is not applicable for that component.*

**Supplementary Figure 1. Photograph of the calibrated obstetric blood collection drape**

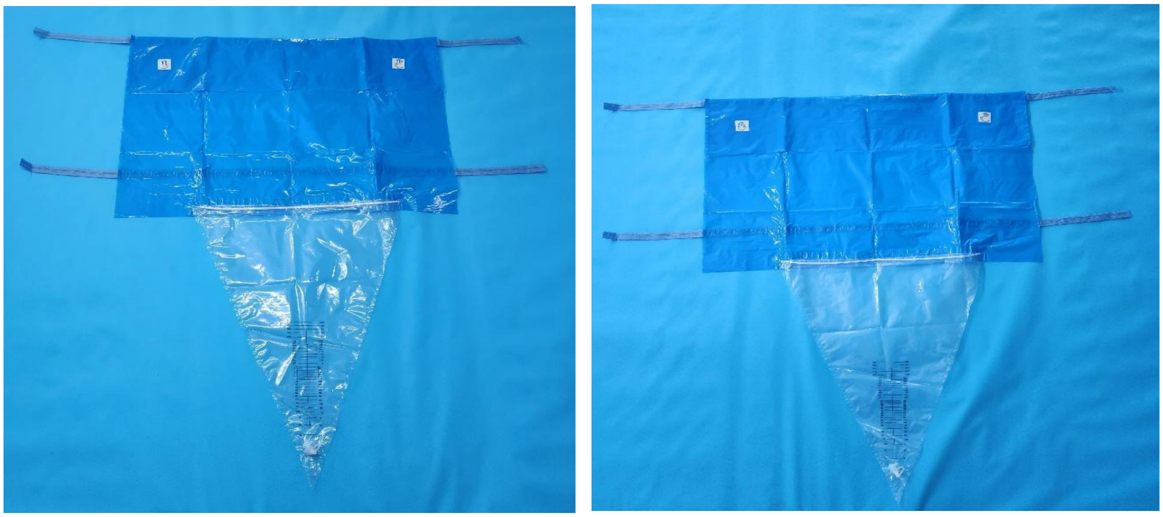

**Pre- and Post-Training Knowledge Assessment**

Thirty participants (15 in each hospital) completed pre- and post-training assessments Baseline knowledge of PPH was generally high across both sites. Correct identification of the standard PPH definition increased from 93% at baseline to 97% post-training, while recognition of clinical risk factors improved from 83% to 93%. Awareness of the importance of early PPH identification remained high before and after training (97%). (**Supplementary Table 2**).

Knowledge related to blood loss measurement methods also showed modest improvements following training. Identification of calibrated obstetric drapes as the most effective method for measuring blood loss increased from 83% to 97%, while recognition of visual estimation as an inaccurate method increased from 70% to 87%. High levels of knowledge regarding infection prevention and drape handling were observed at baseline and were largely maintained following training (**Supplementary Table 2**).

**Supplementary Table 2. Pre- and post-training knowledge assessment: Hospital -1 (n=15) and Hospital -2 (n=15)**

| | **Hospital-1 (n=15)** | | **Hospital-2 (n=15)** | | **Combined (n=30)** | |
|---|---|---|---|---|---|---|
| **Knowledge/practice assessed** | **Pre-test n (%)** | **Post-test n (%)** | **Pre-test n (%)** | **Post-test n (%)** | **Pre-test n (%)** | **Post-test n (%)** |
| Correctly identified the standard definition of PPH | 15 (100%) | 15 (100%) | 13 (87%) | 14 (93%) | 28 (93%) | 29 (97%) |
| Correctly identified visual estimation as inaccurate for blood loss estimation | 12 (80%) | 14 (93%) | 9 (60%) | 12 (80%) | 21 (70%) | 26 (87%) |
| Correctly identified calibrated obstetric drapes as the most effective measurement method | 13 (87%) | 15 (100%) | 12 (80%) | 14 (93%) | 25 (83%) | 29 (97%) |
| Correctly identified recognised clinical risk factors for PPH | 13 (87%) | 15 (100%) | 12 (80%) | 13 (87%) | 25 (83%) | 28 (93%) |

| Knowledge/practice assessed | Hospital-1 (n=15) Pre-test n (%) | Hospital-1 (n=15) Post-test n (%) | Hospital-2 (n=15) Pre-test n (%) | Hospital-2 (n=15) Post-test n (%) | Combined (n=30) Pre-test n (%) | Combined (n=30) Post-test n (%) |
|---|---|---|---|---|---|---|
| Correctly recognised the importance of early PPH identification for timely intervention | 14 (93%) | 15 (100%) | 15 (100%) | 14 (93%) | 29 (97%) | 29 (97%) |
| Correctly identified the primary function of the calibrated drape as accurate blood loss measurement | 15 (100%) | 15 (100%) | 15 (100%) | 14 (93%) | 30 (100%) | 29 (97%) |
| Correctly identified appropriate timing for drape application (before placental delivery)* | 13 (87%) | 15 (100%) | 14 (93%) | 15 (100%) | 27 (90%) | 30 (100%) |
| Correctly identified sterile technique as necessary to prevent cross-contamination | 15 (100%) | 14 (93%) | 15 (100%) | 14 (93%) | 30 (100%) | 28 (93%) |
| Correctly identified reuse after rinsing as an incorrect practice | 12 (80%) | 15 (100%) | 13 (87%) | 14 (93%) | 25 (83%) | 29 (97%) |
| Correctly identified that calibrated drapes must not be reused | 5 (33%) | 0 (0%) | 8 (53%) | 14 (93%) | 13 (43%) | 14 (47%) |
| Correctly identified disposal in a designated biomedical waste bin | 13 (87%) | 15 (100%) | 15 (100%) | 14 (93%) | 28 (93%) | 29 (97%) |
| Correctly identified the need to inspect the drape for damage before use | 14 (93%) | 15 (100%) | 15 (100%) | 14 (93%) | 29 (97%) | 29 (97%) |
| Correctly identified that accurate positioning improves blood loss measurement | 14 (93%) | 15 (100%) | 12 (80%) | 14 (93%) | 26 (87%) | 29 (97%) |
| Correctly identified repositioning using sterile technique as appropriate if drape is displaced | 15 (100%) | 15 (100%) | 9 (60%) | 14 (93%) | 24 (80%) | 29 (97%) |

*Values represent the number (n) and percentage (%) of participants selecting the correct response*
** For this item, responses indicating drape application at the time of fetal delivery or before placental delivery were considered correct, as both represent the appropriate clinical period for drape placement and blood loss collection.*

**Post-Training Outcomes and User Perceptions**

Post-test outcomes were assessed using the highest response category on a five-point Likert scale. Across both sites, 70% of participants reported "very high" confidence in detecting PPH after training, and 80% reported a "very high" understanding of PPH management concepts. Very high perceived effectiveness of the calibrated drape and familiarity with its use were each reported by 80%, while 70% reported very high confidence in correct drape use in practice. Very high clarity of training content was reported by 87% of participants.

Perceptions of the training programme were also positive.: very high effectiveness of practical demonstrations was reported by 97%, usefulness of training materials by 93%, and adequacy of hands-on practice by 90%. Very high overall training satisfaction was reported by 73%, and 83% indicated a high likelihood of using the drape in future practice. Perceived challenges in drape use were acknowledged by 47% of participants. Participants recommended extending training to other staff involved in deliveries (including support workers) and holding periodic refresher sessions (**Supplementary Table 3**).

**Supplementary Table 3. Post-training outcomes and acceptability: Hospital -1 (n=15) and Hospital -2 (n=15)**

| Training outcome assessed | Hospital-1 n (%)* | Hospital-2 n (%)* | Combined n (%) |
|---|---|---|---|
| Confidence in detecting PPH after training | 12 (80%) | 9 (60%) | 21 (70%) |
| Self-reported understanding of PPH management concepts | 14 (93%) | 10 (67%) | 24 (80%) |
| Perceived effectiveness of the calibrated drape for PPH detection | 14 (93%) | 10 (67%) | 24 (80%) |
| Familiarity with using the calibrated obstetric drape | 13 (87%) | 11 (73%) | 24 (80%) |

| **Training outcome assessed** | **Hospital-1 n (%)*** | **Hospital-2 n (%)*** | **Combined n (%)** |
|---|---|---|---|
| Confidence in correct use of the calibrated drape in practice | 13 (87%) | 8 (53%) | 21 (70%) |
| Clarity of training content and instructions | 15 (100%) | 11 (73%) | 26 (87%) |
| Effectiveness of practical demonstrations | 15 (100%) | 14 (93%) | 29 (97%) |
| Usefulness of training materials | 15 (100%) | 13 (87%) | 28 (93%) |
| Adequacy of hands-on practice opportunities | 14 (93%) | 13 (87%) | 27 (90%) |
| Overall satisfaction with the training session | 14 (93%) | 8 (53%) | 22 (73%) |
| Likelihood of using the calibrated drape in future practice | 14 (93%) | 11 (73%) | 25 (83%) |

** Values represent the number (n) and percentage (%) of participants selecting the highest response category (score 5: 'very confident' / 'very effective' / 'very satisfied' / 'very likely') on a five-point Likert scale. PPH, postpartum haemorrhage.*

**On-the-Job Observations**

A total of 20 on-the-job observations were conducted (10 per site). High adherence to recommended practices was observed across both hospitals. Timely drape application, monitoring of blood loss, use of the measurement scale, and decision-making based on measured blood loss were observed in all 20 cases. Correct drape positioning was observed in 90% of cases at Hospital -2 and in 100% of cases at Hospital1.

Some variation was observed in the duration of drape use between sites. At Hospital-2, the drape was typically retained for 10–15 minutes following episiotomy repair and removed once active bleeding had ceased. At Hospital-1, the drape was generally retained until transfer to the postnatal ward following confirmation of haemodynamic stability and cessation of active bleeding. Team involvement and staff familiarity with drape use were consistently observed across both hospitals. (**Supplementary Table 4**).

**Supplementary Table 4. On-the-job observation checklist findings: Hospital -1 (n=10) and Hospital -2 (n=10)**

| Checklist item | Hospital -2 n (%) | Hospital -1 n (%) |
|---|---|---|
| Timing of drape application (prior to delivery of baby) | 10 (100%) | 10 (100%) |
| Correct positioning of drape | 9 (90%) | 10 (100%) |
| Monitoring of blood loss | 10 (100%) | 10 (100%) |
| Decision-making based on measured blood loss | 10 (100%) | 10 (100%) |
| Staff familiarity and confidence with drape | 10 (100%) | 10 (100%) |
| Team involvement in drape use | 10 (100%) | 10 (100%) |
| Use of measurement scale on the drape | 10 (100%) | 10 (100%) |

*n, number of observations; %, percentage of cases in which correct practice was observed.*

**Supplementary Table 5: Standards for Reporting Qualitative Research (SRQR)*(checklist)**

| **Standards for Reporting Qualitative Research (SRQR)*** | **Page/line no(s).** |
|---|---|

| | |
|---|---|
| **Title and abstract** | |
| **Title** - Concise description of the nature and topic of the study Identifying the study as qualitative or indicating the approach (e.g., ethnography, grounded theory) or data collection methods (e.g., interview, focus group) is recommended | Page 1/ Line 1-3 |
| **Abstract** - Summary of key elements of the study using the abstract format of the intended publication; typically includes background, purpose, methods, results, and conclusions | Pages 1/ Lines 7-36 |
| **Introduction** | |
| **Problem formulation** - Description and significance of the problem/phenomenon studied; review of relevant theory and empirical work; problem statement | Pages 2/ Lines 37-58 |
| **Purpose or research questio**n - Purpose of the study and specific objectives or questions | Pages 2/ Lines 49-58 |
| **Methods** | |
| **Qualitative approach and research paradigm** - Qualitative approach (e.g., ethnography, grounded theory, case study, phenomenology, narrative research) and guiding theory if appropriate; identifying the research paradigm (e.g., postpositivist, constructivist/ interpretivist) is also recommended; rationale** | Page 2-3/ Lines 60-70 |
| **Researcher characteristics and reflexivity** - Researchers' characteristics that may influence the research, including personal attributes, qualifications/experience, relationship with participants, assumptions, and/or presuppositions; potential or actual interaction between researchers' characteristics and the research questions, approach, methods, results, and/or transferability | Page 4-5/ Lines 112-127 |
| **Context** - Setting/site and salient contextual factors; rationale** | Page 2-4/ Lines 72-110 |
| **Sampling strategy** - How and why research participants, documents, or events were selected; criteria for deciding when no further sampling was necessary (e.g., sampling saturation); rationale** | Page 4/ Lines 102-110 |
| **Ethical issues pertaining to human subjects** - Documentation of approval by an appropriate ethics review board and participant consent, or explanation for lack thereof; other confidentiality and data security issues | Pages 14/ Lines 405-412 |
| **Data collection methods** - Types of data collected; details of data collection procedures including (as appropriate) start and stop dates of data collection and analysis, iterative process, triangulation of sources/methods, and modification of procedures in response to evolving study findings; rationale** | Pages 4-5/ Lines 112-127 |

| | |
|---|---|
| **Data collection instruments and technologies** - Description of instruments (e.g., interview guides, questionnaires) and devices (e.g., audio recorders) used for data collection; if/how the instrument(s) changed over the course of the study | Pages 4-5/ Lines 112-127 |
| **Units of study** - Number and relevant characteristics of participants, documents, or events included in the study; level of participation (could be reported in results) | Pages 4 & 6/ Lines 102-110 ,152-158 |
| **Data processing** - Methods for processing data prior to and during analysis, including transcription, data entry, data management and security, verification of data integrity, data coding, and anonymization/de-identification of excerpts | Pages 5-6/ Lines 129-148 |
| **Data analysis** - Process by which inferences, themes, etc., were identified and developed, including the researchers involved in data analysis; usually references a specific paradigm or approach; rationale** | Pages 5-6/ Lines 141-148 |
| **Techniques to enhance trustworthiness** - Techniques to enhance trustworthiness and credibility of data analysis (e.g., member checking, audit trail, triangulation); rationale** | Pages 4-5/ Lines 112-127 |

**Results/findings**

| | |
|---|---|
| **Synthesis and interpretation** - Main findings (e.g., interpretations, inferences, and themes); might include development of a theory or model, or integration with prior research or theory | Pages 6-11/ Lines 150-319 |
| **Links to empirical data** - Evidence (e.g., quotes, field notes, text excerpts, photographs) to substantiate analytic findings | Pages 6-11/ Lines 150-319 |

**Discussion**

| | |
|---|---|
| **Integration with prior work, implications, transferability, and contribution(s) to the field** - Short summary of main findings; explanation of how findings and conclusions connect to, support, elaborate on, or challenge conclusions of earlier scholarship; discussion of scope of application/generalizability; identification of unique contribution(s) to scholarship in a discipline or field | Pages 11-13/ Lines 321-388 |
| **Limitations** - Trustworthiness and limitations of findings | Pages 13/Lines 375-388 |

**Other**

| | |
|---|---|
| **Conflicts of interest** - Potential sources of influence or perceived influence on study conduct and conclusions; how these were managed | Pages 14/ Lines 422-423 |
| **Funding** - Sources of funding and other support; role of funders in data collection, interpretation, and reporting | Pages 14/ Lines 425-431 |

*The authors created the SRQR by searching the literature to identify guidelines, reporting standards, and critical appraisal criteria for qualitative research; reviewing the reference lists of retrieved sources; and contacting experts to gain feedback. The SRQR aims to improve the transparency of all aspects of qualitative research by providing clear standards for reporting qualitative research.

**The rationale should briefly discuss the justification for choosing that theory, approach, method, or technique rather than other options available, the assumptions and limitations implicit in those choices, and how those choices influence study conclusions and transferability. As appropriate, the rationale for several items might be discussed together.

**Reference:**

O'Brien BC, Harris IB, Beckman TJ, Reed DA, Cook DA. **Standards for reporting qualitative research: a synthesis of recommendations.** *Academic Medicine*, Vol. 89, No. 9 / Sept 2014
DOI: 10.1097/ACM.0000000000000388

http://www.equator-network.org/reporting-guidelines/srqr/
